# A mobile app providing individually-tailored psychoeducation about sleep for older adults with chronic health conditions and low health literacy

**DOI:** 10.1101/2024.01.02.24300726

**Authors:** Raymond L Ownby, Kamilah Thomas-Purcell, Donrie Purcell, Joshua Caballero, Sweta Tewary, Rosemary Davenport, Michael Simonson

## Abstract

**Objective:** This paper reports the effect of a mobile app that provides tailored information about sleep to individuals aged 40 and older who have chronic health conditions and low health literacy.

**Methods:** The sleep module was a part of a multitopic app focused on chronic disease self-management. Participants were randomly assigned to receive sleep psychoeducation at reading levels equivalent to 3^rd^, 6^th^ or 8^th^ grade. The primary outcome measure was the Pittsburgh Sleep Quality Index (PSQI), which was completed at baseline, after the intervention, and again three months later. Outcomes were assessed using repeated measures mixed effects models.

**Results:** Most participants were Black, Indigenous, or Other Persons of Color (BIPOC; 87%); they had average reading level at the 7^th^ grade. Health literacy, socioeconomic status, number of health conditions, and the intervention’s effect were related to the PSQI. The PSQI score significantly decreased over the course of the three study visits for all groups, consistent with a small to medium effect size (*d* = 0.40). No effect of treatment group was observed.

**Conclusion:** Results suggest that a brief tailored information intervention may be beneficial for individuals aged 40 and older who have low health literacy and chronic health conditions. Further development of the intervention may enhance its clinical effectiveness.

Registered at clinicaltrials.gov NCT02922439.

## Introduction

Sleep problems are common among individuals in the United States and have a significant impact on their mood, quality of life, (National Sleep Foundation, 2023), workplace productivity (Filip et al., 2017), and cognitive functioning (Goel et al., 2009), and risk for chronic diseases (Liu et al., 2013). Despite available treatments, a significant number of individuals do not seek help for sleep disorders (Gordon et al., 2022; Torrens Darder et al., 2021), highlighting the need to prioritize the dissemination of information about the importance of sleep and available treatments (Di et al., 2022).

Among older adults, especially those with chronic medical conditions, sleep disturbances are common and have substantial clinical and public health implications. Onen (2018) found that 83% of older adults report at least one chronic medical condition, and these conditions are associated with higher rates of insomnia, sleep apnea, and other sleep disturbances. Additionally, Foley (2004) noted that conditions such as heart disease, depression, pain, and memory problems are linked to higher rates of insomnia, while obesity, diabetes, and lung disease are linked to sleep apnea, daytime sleepiness, and restless leg syndrome. Nadorff (2018) found that sleep disorders are strongly associated with anxiety, depression, dementia, and suicidal thoughts in older adults.

Sleep problems may be an even greater concern for older adults with chronic health conditions (Liu et al., 2013; Onen & Onen, 2018). The negative impact of sleep problems on daily functioning in older adults has also been reported (Gooneratne & Vitiello, 2014; Manocchia, San, & Ware, 2001), especially among persons from minoritized groups (Meredith et al., 2020). Sleep problems can have a detrimental impact on older adults’ health-related quality of life, daily functioning, and health care utilization (Dai et al., 2019; Manocchia, Keller, & Ware, 2001). Insufficient or poor-quality sleep is associated with chronic health conditions and worse health outcomes, particularly in minoritized and low-income populations (Jackson et al., 2015; Laposky et al., 2016; Williams et al., 2015). Individuals from minoritized racial and ethnic groups and those with lower socioeconomic status (SES) tend to get less sleep and report poorer sleep quality (Jehan et al., 2018; Williams et al., 2015). Sleep problems may contribute to health disparities in conditions such as obesity, cardiovascular disease, and diabetes (Jackson et al., 2015; Laposky et al., 2016; Williams et al., 2015) and various factors, including SES, racism, discrimination, neighborhood segregation, and access to healthcare, can influence sleep health disparities (Billings et al., 2021).

Health literacy, defined as the ability to understand and use health information, plays a crucial role in shaping health outcomes and disparities. Low health literacy has been linked to sleep disturbances (Ghiassi & Partridge, 2011; Hackney et al., 2008) and may be a factor in health disparities (Jackson et al., 2015), as is health literacy itself (Paasche-Orlow & Wolf, 2007, 2010). Persons with lower health literacy tend to be older, less educated, have lower income and report poorer sleep and overall health (Cutilli et al., 2018; Hackney et al., 2008). They may encounter difficulties in understanding and managing sleep disorders or implementing recommendations to improve their sleep health (Hackney et al., 2008). Several studies have demonstrated a link between inadequate health literacy and poor sleep health knowledge or outcomes. Hackney (2008) reviewed research showing patients with low health literacy understand less about sleep disorders and engage in poorer sleep hygiene. Gazmararian (2003) found that patients with chronic diseases and low health literacy have less knowledge about managing their conditions. Morrow (2006) found that adults with chronic heart failure and lower health literacy tended to have worse sleep, cognition, and more health problems, suggesting that health literacy interventions should consider patients’ sleep and cognitive abilities. A complex group of interactions between sleep, chronic disease, and health literacy may thus exist.

Treatment options for sleep disorders in older adults encompass a range of approaches, both pharmacological and non-pharmacological (Bloom et al., 2009). Non-drug therapies particularly cognitive behavioral therapy can be highly effective and have long-lasting benefits (Gooneratne & Vitiello, 2014). Medications may also be used but can cause side effects such as impaired thinking or movement (Gooneratne & Vitiello, 2014). Given the bidirectional relationship between sleep and health in older adults, accurate diagnosis and proper treatment of sleep disorders are crucial.

Evidence suggests that older adults often rely on pharmacologic treatments such as over-the-counter sleep aids or prescription sleep medications (Abraham et al., 2017; Musich et al., 2018), highlighting the need for accessible alternatives such as digital interventions. Although behavioral treatments have demonstrated effectiveness, access to them is limited by the number of trained providers and their distribution (Thomas et al., 2016). One strategy to address these problems is the adaptation of behavioral sleep medicine techniques to digital interventions making them more widely available. Several apps, for example, for cognitive behavioral therapy for insomnia (CBT-I), have shown promise in helping people improve their sleep (Espie et al., 2016; Kuhn et al., 2016; Yu et al., 2018).

Digital therapeutics and mobile health interventions show great potential for expanding healthcare access and reducing health disparities. Rivers (2014) argues that mobile technology and mHealth interventions can play a vital role in addressing health disparities by providing accessible health information and care to underserved groups. Similarly, Gibbons (2011) proposes that Web 2.0 technologies such as social media could be harnessed to provide health promotion and education to minoritized and medically underserved groups. Several authors have emphasized the need to address sleep disparities and proposed actionable strategies (Billings et al., 2021; Hughes et al., 2022; Jackson et al., 2020; Miller et al., 2023). By applying health equity frameworks, engaging communities, and targeting social determinants of health, digital therapeutics can fulfil their potential to reduce disparities and empower marginalized groups.

This paper presents the results of a study of a multitopic mobile app for chronic disease self-management (CDSM). The app included a module providing individually-tailored psychoeducation on sleep to older adults with chronic health conditions and low health literacy. Participants’ response to the app and the relations of sleep to their SES, health literacy, and chronic health conditions, as well as the effects of the intervention, were evaluated.

## Method

The app’s content was initially developed by examining existing resources related to CDSM and conducting a qualitative study to identify the information needs of older adults with chronic health conditions (Jacobs et al., 2017; Thomas-Purcell et al., 2019). Input from potential users was crucial in shaping the content and format of the intervention. To ensure evidence-based content, a diverse team was assembled that comprised professionals from relevant disciplines including medicine, nursing, psychology, pharmacy, public health, and education.

The app consisted of thematic sections, each presenting content on multiple screens. Guided by cognitive load theory-based instructional design (Paas et al., 2004), each module included an introduction, an assessment of participants’ current status, general health-related insights, personalized content, and a summary. Participants could test their understanding through self-check questions. Information was presented through text complemented by images, diagrams, and narrated animations in line with the principles of multimedia learning (Mayer, 2009).

More detailed information on the app’s initial development and testing can be found in a recently-published paper (Patel et al., 2023). The modules were devised with identical content but different reading complexity levels, categorized based on the Fry and Flesch Reading Ease scores (3rd grade, with narration; 6th and 8th grade). This was facilitated through the use of Health Literacy Advisor® software integrated with Microsoft Word®.

The study took place at two locations: Fort Lauderdale, Florida and Atlanta, Georgia. Participants were recruited through various channels, such as past unrelated studies, local healthcare clinics, medical practices, and word of mouth. In Atlanta, collaboration with local churches helped identify and inform potential participants about the study. Data collection followed U.S. National Institutes of Health guidelines, including recording participant information including race, ethnicity, and gender.

Screening began with a brief telephone interview to determine eligibility based on medical history, medication use, and educational background. The Rapid Estimate of Adult Literacy in Medicine was used with a predefined cutoff for health literacy set at or below an 8th-grade level (Han et al., 2017, October). Eligibility criteria included being 40 years or older, having at least one ongoing chronic health condition under treatment, having an education below college graduate, and a score below the cutoff on the health literacy assessment. The college education criterion stemmed from prior research showing that successful completion of a college degree correlated with adequate health literacy (Ownby et al., 2019).

Participant assessments were completed at baseline, post-intervention, and at a follow-up three months later. The FLIGHT/VIDAS health literacy scale (Ownby, Acevedo, Jacobs, et al., 2014; Ownby et al., 2013; Ownby, Acevedo, Waldrop-Valverde, et al., 2014; Ownby et al., 2021) was chosen due to its comprehensive score range, unlike to other measures with ceiling effects (Pleasant, 2014). Participants’ socioeconomic status was evaluated using an index created through a principal components analysis of self-reported education and income; component scores were used for data analysis.

Participants’ health status was assessed through a structured health conditions interview based on the Functional Comorbidity Scale (Groll et al., 2005) but expanded to include additional health conditions common in older adults (Centers for Medicare and Medicaid Services, 2012). Self-report measures were administered via audio computer-assisted self-interview software (Bethesda MD: Questionnaire Development System) that read all questions aloud to participants to minimize the effect of reading ability on their responses.

### The sleep module

Table 1 provides an outline of the topics covered in the module. After viewing a section on the purpose of the module, participants completed the Insomnia Severity Index or ISI (Morin et al., 2011). Responses to the ISI were used to individually tailor content presented later in the module. For example, participants who reported difficulty in getting to sleep received suggestions to address it. Next, participants received feedback on how depression and stress could potentially impact their sleep based on their responses to the CES-D and PSS. The module then proceeded to discuss sleep stages, the two-process theory (Borbély, 1982), and how chronic health conditions can affect sleep. It continued with a review of sleep hygiene, approaches for keeping a sleep log, and a discussion of sleep restriction.

**Table 1.** Content overview of chronic disease self-management sleep module.

| Topics |
| --- |
| Purpose of the module |
| Association of chronic health conditions and sleep problems |
| Association of sleep disturbance with depressive symptoms and stress |
| Assessment of current sleep status via Insomnia Severity Index |
| Personalized feedback on participants' levels of depressive symptoms and stress from the CES-D and PSS |
| Sleep and the immune system |
| What is sleep? Understanding that it's a time when the brain is active and that good sleep is essential for functioning |
| Basic information about sleep, including sleep stages and importance of each, with narrated animations illustrating the stages and explaining movement from one stage to the next. |
| How chronic health conditions can affect sleep via factors such as pain and shortness of breath. |
| The two process model of sleep, linking the model to factors likely to improve or adversely affect sleep |
| Rules for better sleep: Sleep hygiene recommendations |
| How to use a sleep diary to see patterns related to medication and substance use, daily activity, and stress. |
| Feedback on responses to the Insomnia Severity Index with individualized recommendations to address problems reported. |
| Creating an action plan to improve sleep |
| Summary |

### Statistical analyses

Statistical analyses assessed the impact of SES, health literacy, number of health conditions, the intervention and treatment group assignment. Data processing and analysis were performed using SPSS and R (R Core Team, 2022). The intended sample size was determined using the mixed effects model simulation routine in PASS (NCSS, 2018), ensuring a minimum of 30 participants per group for adequate statistical power.

The effects of covariates and the intervention were evaluated using a series of random intercept mixed effects models that progressively increased in complexity (Luke, 2017). These were implemented with the R package *lme4* (Bates et al., 2015). The order in which covariates were included in the models was determined based on their anticipated impact on outcomes and potential for intervention. The baseline model (model 1 in Table 3) included only age, race, and gender (model 1). Subsequently, study site, SES, health literacy, health conditions, intervention effect over time, and treatment group were added. After computing model 1, likelihood ratio tests were conducted to compare the previous model to one with an additional covariate in order to assess its significance. To facilitate interpretation of results, chi-square values obtained from model comparisons were converted to Cohen’s *d* (J. Cohen, 1988) using the R package *esc* (Ludecke, 2019).

### Human subjects approval

All procedures were approved by the Institutional Review Boards of Nova Southeastern University (2018-685-NSU) and of Emory University (MODCR001-IRB00087112). Participants provided verbal consent for initial screening procedures and written informed consent for all other aspects of their participation.

## Results

A total of 286 individuals completed the baseline assessments and the sleep module. Of these individuals, 195 had ISI scores of 8 or greater, suggesting insomnia according to the criterion reported by Morin et al. (2011). This report focuses on the data from these participants. Data from these participants are reported here. Of the 195 participants, 91 (46.7%) were men and 104 (53.3%) were women. There were 168 participants who identified as Black, Indigenous, or Other People of Color (BIPOC) accounting for 86% of the sample, while 27 (14.0%) identified as white individuals. The majority (61%) reported annual incomes of less than $10,000 (US). Following the interpretation suggested by Morin et al. (2011), 87 (45%) had subthreshold, 79 (41%) had moderate, and 29 (15%) had severe insomnia. Complete information on recruitment, inclusion, and randomization, including a CONSORT diagram detailing patient flow through the study, is available online as a preprint (Ownby et al., 2023).

Descriptive statistics for continuous variables are presented in Table 2. The average level of health literacy on the FLIGHT/VIDAS measure was 10.02, indicating a basic level of health literacy that would enable a person to manage simple tasks such as reading straightforward prescription labels (Ownby et al., 2019). However, persons at this level of health literacy may struggle with understanding more complex directions or text that requires inferences. Their average score on the PSS was at a level above the population averages previously reported (S. Cohen & Janicki-Deverts, 2012), and their average score on the CES-D was greater than typical cutoff scores for major depression (Vilagut et al., 2016). Average scores on the PSQI and ISI were consistent with a substantial sleep disturbance. Participants spent an average of 32 minutes completing the sleep module (SD = 13.7).

**Table 2.**
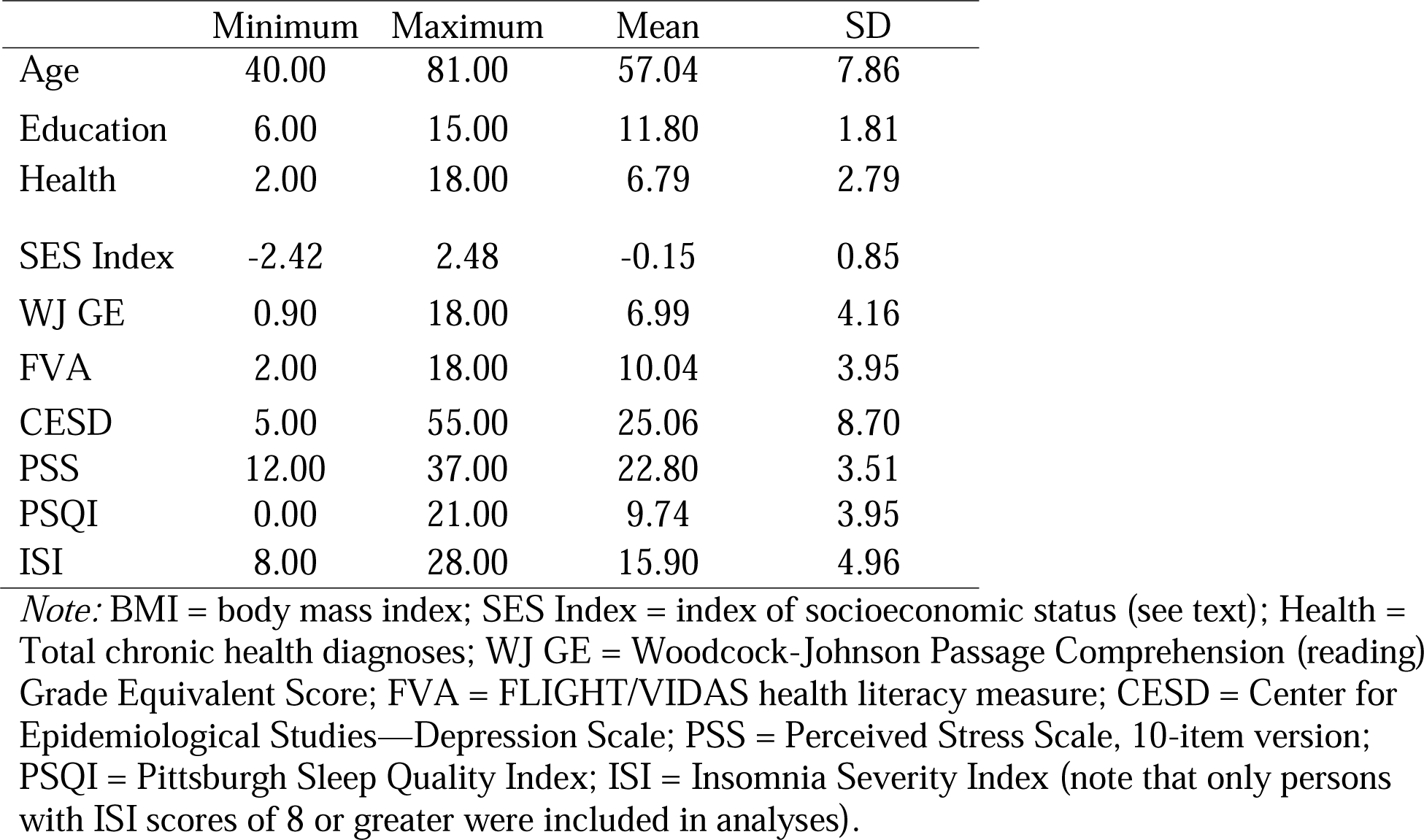
Descriptive statistics for continuous variables (n = 195)

|  | Minimum | Maximum | Mean | SD |
| --- | --- | --- | --- | --- |
| Age | 40.00 | 81.00 | 57.04 | 7.86 |
| Education | 6.00 | 15.00 | 11.80 | 1.81 |
| Health | 2.00 | 18.00 | 6.79 | 2.79 |
| SES Index | -2.42 | 2.48 | -0.15 | 0.85 |
| WJ GE | 0.90 | 18.00 | 6.99 | 4.16 |
| FVA | 2.00 | 18.00 | 10.04 | 3.95 |
| CESD | 5.00 | 55.00 | 25.06 | 8.70 |
| PSS | 12.00 | 37.00 | 22.80 | 3.51 |
| PSQI | 0.00 | 21.00 | 9.74 | 3.95 |
| ISI | 8.00 | 28.00 | 15.90 | 4.96 |

*Note:* BMI = body mass index; SES Index = index of socioeconomic status (see text); Health = Total chronic health diagnoses; WJ GE = Woodcock-Johnson Passage Comprehension (reading) Grade Equivalent Score; FVA = FLIGHT/VIDAS health literacy measure; CESD = Center for Epidemiological Studies—Depression Scale; PSS = Perceived Stress Scale, 10-item version; PSQI = Pittsburgh Sleep Quality Index; ISI = Insomnia Severity Index (note that only persons with ISI scores of 8 or greater were included in analyses).

### Chronic health conditions and sleep disturbance

We evaluated which health conditions might be related to sleep disturbance by assessing the presence or absence of a condition related to sleep disturbance defined as having a PSQI score greater than 5 (Buysse et al., 2008) in multiple chi-square analyses uncorrected for multiple comparisons. The relation of sleep disturbance to chronic obstructive pulmonary disease approached statistical significance (*χ*^2^ [1] = 3.53, *p* = 0.06) as did that for angina (*χ*^2^ [1] = 3.57, *p* = 0.06). It was related to a diagnosis of hypercholesterolemia (*χ*^2^ [1] = 5.10, *p* = 0.02), depression *χ*^2^ [1] = 10.60, *p* = 0.001), other nervous condition (*χ*^2^ [1] = 13.85, *p* = < 0.001), and bipolar disorder (*χ*^2^ [1] = 6.77, *p* = 0.009).

### Models for the PSQI

Results of the likelihood ratio tests for sequential assessments of progressively more complex models for the PSQI are presented in Table 3. These models represent, in order, baseline (including age, gender, and race as covariates), site (Fort Lauderdale vs Atlanta), SES (an index of education and income), health literacy, total health conditions, time (baseline, immediate follow-up, and three-month follow-up), and treatment group (3^rd^, 6^th^, or 8^th^ grade reading level). It can be seen that multiple factors are significantly associated with sleep quality over time, including SES, health literacy, and total health conditions. The effect of the intervention (assessed change in PSQI over time) was statistically significant and consistent with a small to medium effect size (*d* = 0.40). Figure 1 displays a plot of the model-estimated means for each group during the three study visits. This model-corrected change illustrated in the figure represents a decline in PSQI of 0.85 points (95%CI [0.11, 1.60]).

**Table 3.** Likelihood ratio tests for progressively more complex models in random intercept analysis.

| | Parameters | Log<br>Likelihood | $\chi^2$ | *df* | *p* | *d* | 95% CI | Effect |
| --- | --- | --- | --- | --- | --- | --- | --- | --- |
| model1 | 6 | -1270 |  |  |  |  |  | Base |
| model2 | 7 | -1270 | 0.62 | 1 | 0.43 | 0.11 | -0.17, 0.39 | Site |
| model3 | 8 | -1270 | 0.01 | 1 | 0.93 | 0.01 | -0.27, 0.80 | SES |
| model4 | 9 | -1264 | 12.01 | 1 | 0.001 | 0.51 | 0.22, 0.80 | HL |
| model5 | 10 | -1260 | 6.84 | 1 | 0.01 | 0.38 | 0.10, 0.67 | Health |
| model6 | 12 | -1257 | 7.46 | 2 | 0.02 | 0.40 | 0.11, 0.69 | Time |
| model7 | 14 | -1256 | 0.56 | 2 | 0.76 | 0.11 | -0.17, 0.39 | Group |

*Note:* For all models, the Pittsburgh Sleep Quality Index in the dependent variable, measured at three time points. Model 1 is base model that included only age, gender and race; Model 2 added the effect of site (Atlanta vs. Fort Lauderdale) to model 1; model 3 added socioeconomic status (SES) to model 3; model 4 added health literacy to model 3; model 5 added total number of health conditions; model 6 added the time (the effect of the intervention over study visits); and model 7 added the effect of treatment group (whether participants received the intervention at 3<sup>rd</sup>, 6<sup>th</sup>, of 8<sup>th</sup> grade reading difficulties).

**Figure 1.**
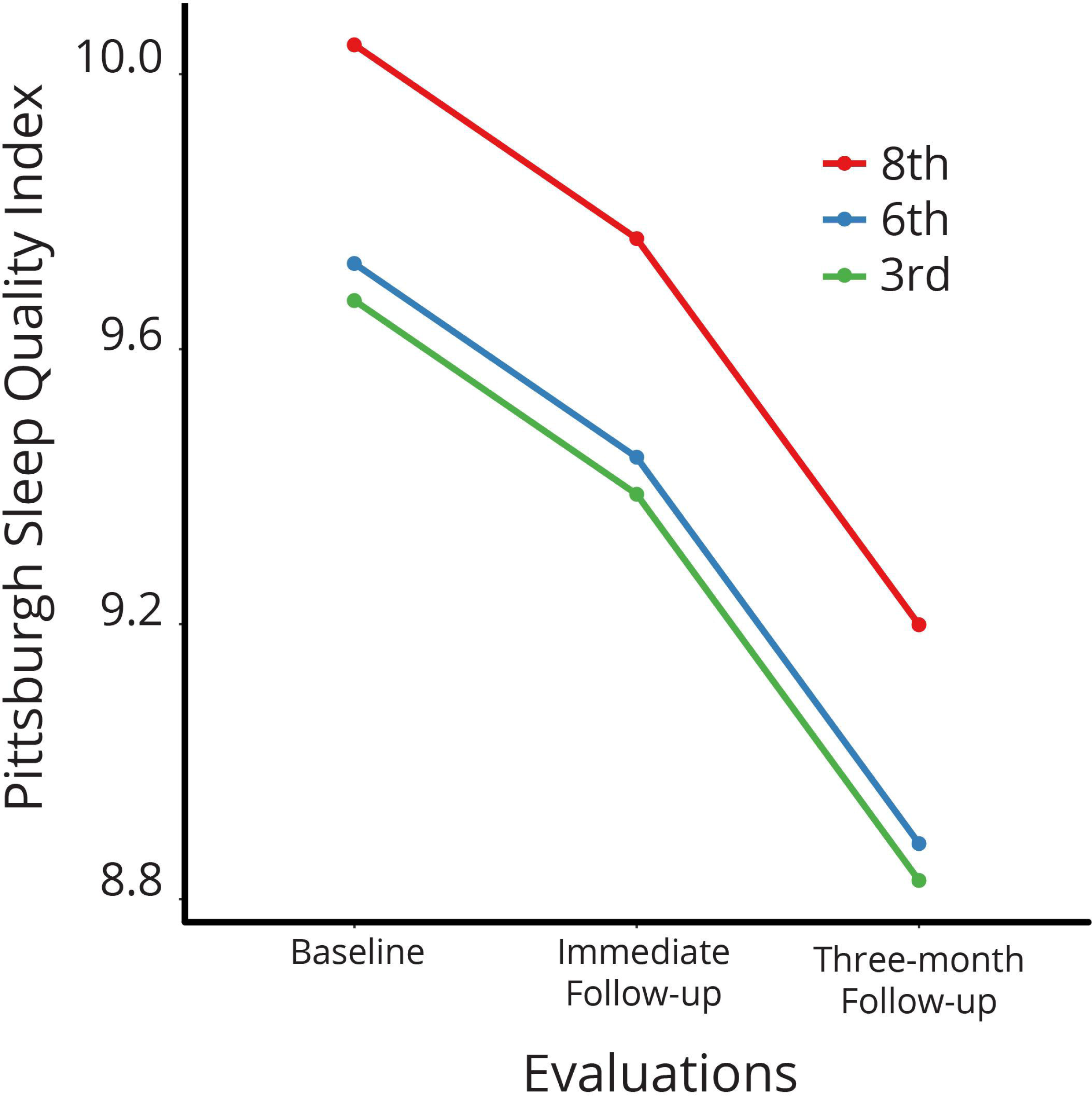
Pittsburgh Sleep Quality Index at Each Visit for Treatment Groups. Figure note: PSQI = Pittsburgh Sleep Quality Index; Evaluations = Study Visit ( 0 = Baseline; 1= First Follow-Up; 2 = Second Follow-Up; Colors represent model-corrected mean values for each treatment group (3^rd^, 6^th^, and 8^th^ grade reading difficulty of intervention materials).

## Discussion

The purpose of this paper was to report on the effects of a brief tailored information intervention for sleep psychoeducation. The intervention specifically targeted individuals aged 40 years and older with chronic health conditions and low health literacy. The sleep module was part of a larger intervention for chronic disease self-management which was delivered in three sessions over two to three weeks. Results of tests of the primary hypotheses of the main study are currently under review and available online as a preprint (Ownby et al., 2023). In addition, we assessed the impact of individual topic modules on relevant measures, and in this instance, we evaluated the modules’ impact on sleep using the PSQI. We found significant decreases in participants’ PSQI scores over the course of the study. While the magnitude of the change was relatively small, it may still be important considering the brevity of the intervention and the limited resources available to individuals with limited economic resources and low health literacy.

Several studies have examined the impact of CBT-I interventions on sleep in older adults. One study (Riedel et al., 1995) found that watching a 15-minute educational video had a significant positive effect on sleep satisfaction compared to a control condition. Participants in this study were older adults with insomnia with an average age of 67.4 years. Some participants in this study were excluded due to chronic health conditions and psychotropic medication use. In another study Lichstein et al. (2001), individuals aged 60 years or older participated in multisession training in either relaxation or sleep compression strategies. This study showed positive effects on sleep satisfaction, with moderate effect sizes. Participants with high levels of caffeine, nicotine, and alcohol use as well as those with elevated levels of anxiety and depression, were excluded from this study.

Buysse et al. (2011), on the other hand, included individuals who might be considered as having comorbid insomnia related to medical or psychiatric disorders, groups previously excluded in some studies. They found that a brief behavioral intervention delivered in two sessions with two follow-up phone calls had a significant effect compared to an information-only control group. The effect size for this intervention was large (*d* = 1.10). It should be noted that the present study’s exclusion criteria were different from those of these studies. Some studies excluded individuals with common comorbidities and substance or medication use, while in our study these individuals participated. Participants’ sleep in our study was less rigorously evaluated than in these studies, but at the same time may have been more representative of individuals who might not have participated in those studies.

Other researchers have explored the effectiveness of brief interventions for sleep problems. Reid et al. did not find an effect on the PSQI of one session of sleep hygiene education (2010), but Tucker et al. (2021) reported a large effect size (*d* = 0.77) for an online six-session educational intervention. In a study that examined the optimal number of sessions in interventions for insomnia, Edinger et al. (Edinger et al., 2007) found that one session was effective in the short term but did not produce lasting effects. Okun and Glidewell (2023) showed that a single four-hour group class was effective in reducing insomnia symptoms in a group whose age averaged 54 years and who had multiple health conditions. These studies suggest that brief interventions can have positive effects on sleep, but longer interventions may have greater and longer-lasting impacts. Despite the smaller effect size observed in our study compared to others, the intervention’s online availability and relatively brief nature may make it a useful initial element of a stepped-care approach (Espie, 2009) for sleep problems. Individuals who do not see improvement after the brief intervention can be referred for more intensive treatment.

The limitations of the present analyses should be acknowledged. The primary focus of the study was chronic disease self-management, with the sleep intervention being just one aspect of it. Participants also completed tailored information modules for mood disturbance, pain, and stress which may have influenced their perception of sleep. Additionally, it is important to note that this report relies on a single self-report measure of sleep, leaving unanswered the question of how the intervention might have affected objective sleep measures. Further, our participants were primarily individuals with low incomes and minoritized groups, limiting the generalizability of these findings to others. Finally, although the intervention did have a statistically significant effect on the PSQI, the change represented a small to medium effect size. It is likely that a more intensive intervention could yield more substantial and clinically meaningful improvements in sleep.

In summary, this paper aimed to present the sleep-specific outcomes of an intervention targeting a range of chronic disease-related issues in individuals aged 40 and older with chronic health conditions and low health literacy. Results suggest a modest improvement in participants’ sleep over the course of the study and follow-up assessments. These findings suggest that a brief intervention offering personalized sleep information and guidance at appropriate reading levels may be helpful in improving sleep. Further development of the intervention appears warranted.

## Data availability

Data from this study are available on request from the first author.

